# The Differential Burden of Acute Rhinovirus Infections in Children with Underlying Conditions

**DOI:** 10.1101/2024.10.23.24315981

**Authors:** María Isabel Sánchez Códez, Isabel Benavente Fernández, Katherine Moyer, Amy L. Leber, Octavio Ramilo, Asuncion Mejias

**Author notes:** **Correspondence:** Asuncion Mejias, MD, PhD, MsCS, St Jude Children’s Research Hospital, 262 Danny Thomas Pl, Mail Stop 320, Memphis, TN, 38105. **Ethical statement:** The study was approved by Nationwide Children’s Hospital (NCH) Institutional Review Board (IRB ID-STUDY00001015). **Authors contributions:** MISC: Study design, writing-original draft, revisions, and data analyses. KM: data collection and interpretation, manuscript review. IBF: data curation and analyses and interpretation and manuscript review. ALL: providing virology data, data interpretation, manuscript review. OR: data interpretation and manuscript review. AM: Study design, data collection and analysis, data interpretation, manuscript review. All authors have read and agreed with the revised version of the manuscript.

## Abstract

**Introduction:** Rhinoviruses (RVs) are well-known trigger of wheezing episodes in children with asthma. Their role in other pediatric chronic medical conditions is not fully know.

**Methods:** Patients ≤21 years hospitalized or evaluated as outpatients with symptomatic RV infection were identified from 2011-2013. Patients were categorized based on the type of underlying disease and differences in clinical parameters, RV loads (C_T_ values), viral and bacterial coinfections and clinical outcomes compared between groups. Multivariable analyses were performed to identify the comorbidities associated with oxygen requirement, PICU admission, and prolonged hospitalization.

**Results:** Of 1,899 children analyzed, 77.7% (n=1477) had an underlying comorbidity including asthma (36.8%), prematurity (7.7%), chronic respiratory diseases (6.4%), congenital heart disease (CHD, 3.2%), immunocompromised hosts (ICH; 1.4%) and others (22.2%). Prevalence of comorbidities increased with age (70%, infants *vs* 84%-87%, children >1 year; p<0.0001). Median RV loads were intermediate-high (24-26 C_T_ values), irrespective of the underlying disease. RV/ viral co-detections were identified in 11% of ICH vs 20%-30% in all other children while bacterial co-infections were identified in 2.9% of children. Multivariable models identified asthma, prematurity, CHD and bacterial coinfections consistently associated with all three clinical outcomes (p<0.0001). Older age and higher RV loads were also associated with increased odds of PICU admission.

**Conclusions:** The prevalence of comorbidities was high in children with RV infections. Of those, asthma, prematurity and CHD were consistently associated with severe disease. Bacterial co-infections and higher RV loads further predicted worse clinical outcomes, highlighting the importance of identifying clinical phenotypes for future targeted interventions.

## INTRODUCTION

Rhinoviruses (RVs) are a leading cause of upper and lower respiratory tract infections (LRTI) in children and adults ^1,2^. The causative role of RVs on respiratory diseases has been underestimated as they are commonly detected in asymptomatic subjects and associated with prolonged shedding ^3-5^. Nonetheless, RVs have been recognized as important triggers of wheezing episodes in asthmatic children ^6,7^. In addition to asthma, pediatric studies suggest that certain underlying conditions, such as congenital heart disease (CHD), prematurity, or immunocompromised hosts (ICH) have an increased risk for severe acute RV infections (ARI) ^1,8-11^. Whether the increased risk for severe RV disease extends to children with other comorbidities, and whether specific clinical parameters or co-detection with other pathogens are associated with increased morbidity is not fully known.

The aim of the present study was to define the clinical phenotype and the differences in clinical presentation, healthcare utilization and the impact of RV loads and of viral/bacterial coinfections in a large cohort of pediatric patients with RVs infection according to the type of underlying condition.

## MATERIALS AND METHODS

### Patient population and study design

Retrospective study of children and adolescents ≤21 years of age with symptomatic RVs infection diagnosed as inpatients or outpatients from 7/2011 through 12/ 2013. Children were identified by virology reports, their clinical was data extracted from electronic healthcare records (EHR), and manually reviewed by two independent investigators. Children with incomplete data and non-respiratory diagnoses were excluded. Duplicate encounters during the same calendar year were also excluded. The present study involves the analysis of a subset of children as part of a larger cohort that has been previously published ^12^.

RV testing was performed per standard of care in respiratory samples using a RV rt-PCR targeting the 5 ′NCR (or 5′UTR) region as described ^13,14^. RV molecular typing was not performed. Semiquantitative RV viral loads were reported as cycle threshold (C_T_) values, with a C_T_ value > 40 being considered not detected. RV testing was run in parallel with individual PCRs for respiratory syncytial virus (RSV), parainfluenza virus (PIV), influenza virus A/B, adenovirus (ADV), and human metapneumovirus (hMPV) as part of a panel ^15^. EHR were assessed for pertinent demographic and clinical data including the admission diagnosis, laboratory and radiologic findings and co-infections. Viral co-detections were defined as the simultaneous detection of one the viruses included in the panel, and bacterial coinfections as the isolation of a bacterial pathogen by culture from a sterile site (blood, pleural fluid), the lower respiratory tract or a throat culture in symptomatic children with cystic fibrosis (CF). We collected information regarding healthcare utilization including medications, need for hospitalization, supplemental oxygen, PICU care and total length of stay. For analyses purposes children were grouped into seven categories based on the type of underlying disease: asthma; other chronic respiratory conditions; prematurity without chronic lung disease (CLD); CHD; ICH; others (i.e endocrine, renal, etc); and (7) none (previously healthy). For children with more than one comorbidity, the most relevant at the time of testing was selected, such as a child with asthma and hypertension was included in the asthma cohort.

The study was approved by Nationwide Children’s Hospital (NCH) Institutional Review Board (IRB ID-STUDY00001015). It was granted exemption from obtaining informed consent due to its retrospective design in accordance with the US Department of Health and Human Services regulations, specifically 45 CFR 46. Data was accessed from EHR over a 12-month period (January 6^th^ through December 19^th^ of 2014), and any identifier linked to participant individuals destroyed upon data collection and curation.

### Statistical Analysis

Qualitative data were expressed as percentages, and quantitative variables as medians and 25%-75% interquartile range (IQR) as data was not normally distributed. Chi-squared test or Fisher’s exact tests were used to compare proportions, and Kruskal–Wallis tests for continuous data. Multivariable logistic regression was performed to define the risk for supplemental oxygen, PICU admission and longer duration of hospitalization according to the type of comorbidity, adjusted by age, sex, RV loads and viral and bacterial coinfections. Two-sided p-values <0.05 were considered statistically significant. Analyses were conducted using STATA 16.0. (StataCorp, College Station, TX, USA).

## RESULTS

### Demographic and Clinical Characteristics

Of the 1,899 children with symptomatic RV infection included, 77.7% (n=1,477) had underlying comorbidities. Asthma was the most prevalent (36.8%; 699) followed by prematurity (7.7%; 147); other chronic respiratory diseases (6.4%; 121); CHD (3.2%; 61); and ICH (1.4%; 27). The remaining children with other comorbidities (22.2%; 422) included genetic syndromes, gastrointestinal, neurologic, hematologic, or endocrine abnormalities (**Table S1**).

Median age for children with comorbidities (15.8 [4.9-52.9] months) was greater than for previously healthy (4.7 [1.5-19.9] months, p=0.0001); **Table 1**.

**Table 1.** Demographics, laboratory, radiology parameters and distribution of diagnoses of children and adolescents with RV infection*.

|  | Previously healthy<br>(n=422; 22.2%) | Asthma/Atopy<br>(n=699; 37%) | Chronic resp conditions †<br>(n=121; 6.4%) | Prematurity<br>(n= 147; 7.7%) | CHD<br>(n=61; 3.2%) | ICH<br>(n=27; 1.4%) | Others §<br>(n=422; 22.2%) | p value |
| --- | --- | --- | --- | --- | --- | --- | --- | --- |
| **Demographic characteristics** |  |  |  |  |  |  |  |  |
| Age, (months) | 4.7 [1.5-19.9] | 23 [8.2-68.5] | 11.6 [5.5-21.2] | 6.7 [2.6-18.7] | 14.7 [3.5-53.4] | 68.6 [42.5-116.3] | 10.5 [3.1-36.8] | **0.0001** |
| Sex, n (%) male | 243 (57.6) | 416 (59.5) | 65 (53.7) | 87 (59.2) | 44 (72.1) | 19 (70.4) | 263 (62.3) | 0.1737 |
| **Laboratory/Radiology data** |  |  |  |  |  |  |  |  |
| WBC (cells/ul) | 10,900 [8,400-14,900] | 13,500 [9,600-17,200] | 11,700 [8,600-15,700] | 11,100 [7,900-16,000] | 10,600 [8,900-16,700] | 2,000 [1,400-3,600] | 11,200 [7,900-16,500] | **0.0001** |
| Lymphocytes (%) | 39 [20-59] | 15 [8-34] | 25 [19-47] | 36.5 [20-52] | 36 [8-59] | 42 [25-63] | 49 [29-65] | **0.0001** |
| ANC (x10cells/ul) | 5,100 [2,700-9,700] | 10,700 [6,900-15,200] | 7,600 [4,300-11,300] | 6,100 [11,900-12,500] | 6,900 [4,200-13,800] | 2,100 [300-3,900] | 6,100 [2,900-11,500] | **0.0001** |
| Chest X-ray performed, n (%) | 236 (55.9) | 541 (77.4) | 94 (77.7) | 111 (75.5) | 49 (80.3) | 16 (59.3) | 289 (68.5) | **0.0001** |
| Chest X-ray abnormalities, n (%) | 183/236 (77.5) | 445/541 (82.2) | 81/94 (86.2) | 83/111 (74.8) | 41/49 (83.7) | 8/16 (50.0) | 233/289 (80.6) | **0.0170** |
| **Diagnosis** |  |  |  |  |  |  |  |  |
| URI | 204 (48.3) | 119 (17.0) | 14 (11.6) | 35 (23.8) | 19 (31.1) | 15 (55.6) | 188 (44.5) | **0.0001** |
| Fever | 8 (1.9) | 9 (1.3) | 0 (-) | 2 (1.4) | 2 (3.3) | 1(3.7) | 7 (1.7) | 0.6436 |
| AOM/sinusitis | 9 (2.1) | 3 (0.4) | 0 (-) | 1 (0.7) | 0 (-) | 1 (3.7) | 8 (1.9) | 0.9985 |
| Wheezing illnesses | 102 (24.2) | 443 (63.4) | 51 (42.2) | 73 (49.7) | 22 (36.1) | 7 (25.9) | 116 (27.5) | **0.0001** |
| PNA/LRTI | 78 (18.5) | 94 (13.5) | 51 (42.2) | 21 (14.3) | 14 (23.0) | 2 (7.4) | 72 (17.1) | **0.0001** |
| Respiratory Failure | 21 (4.9) | 31 (4.4) | 5 (4.1) | 15 (10.2) | 4 (6.6) | 1(3.7) | 31 (7.3) | 0.0877 |

\*Continuous variables are expressed as medians 25–75% interquartile ranges (IQR) and categorical data as frequency (%). Continuous variables were analyzed by Kruskal-Wallis test followed by Holm post-hoc test. Categorical data were analyzed by $\chi^2$ test. Numbers in bold indicate significant p values. ANC, absolute neutrophil count; AOM, acute otitis media; CHD, congenital heart diseases; ICH, immunocompromised host; LRTI, lower respiratory tract infection; PNA, pneumonia; URI, upper respiratory infection; WBC: white blood cell.
†Chronic respiratory conditions included: cystic fibrosis (n=18); chronic lung disease (CLD) (n=61); tracheoesophageal fistula, pulmonary hypertension, spontaneous pneumothorax, airway malacia, subglottic stenosis, cleft palate, and Pierre–Robin sequence. § Others included: genetic syndromes (n=132); gastrointestinal diseases (n=68); neurological disorders (n=36); hematologic disease (n=17) and varied including endocrine, osteoarticular, and genitourinary underlying diseases (n=169).

ICH were the oldest group (68.6 [42.5-116.3] months). The prevalence of comorbidities increased after the first year of life, where 84% to 87% of children had a chronic medical condition vs 70% in infants (p<0.0001; **Fig 1**).

**Figure 1.**
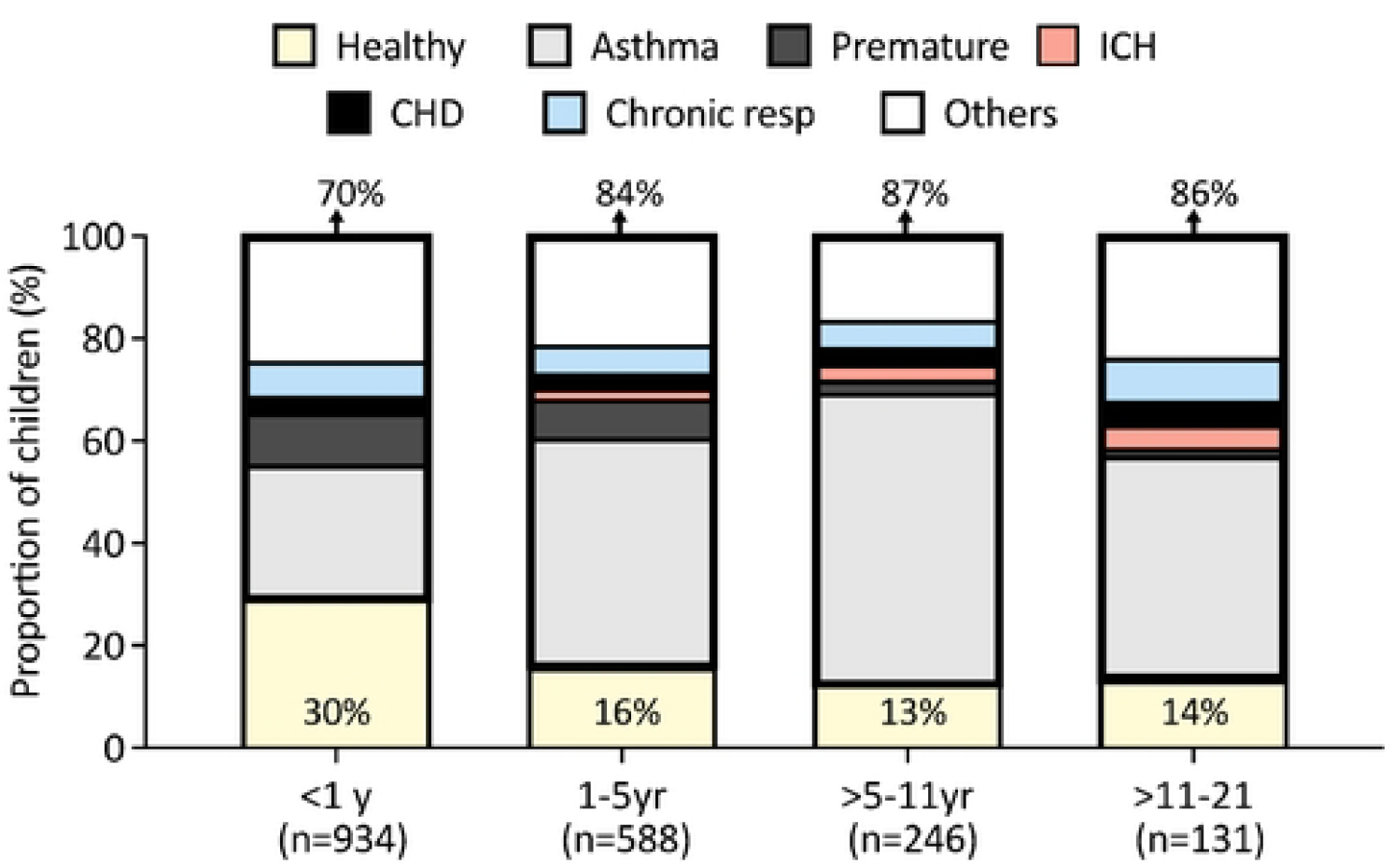
Distribution of underlying conditions according to age. The Y axis depict the proportion of children and adolescents with RV ARI and the X axis the different age groups, specifying the number included in each group underneath the bars. The percentages included at the top of the barrs indicate the proportion of children with underlying conditions for that age group, and those included at the bottom the proportion of previously healthy children per age group, that are color-coded in yellow. The different underlying comorbidities are color coded in black, grey, white or red. ICH, immunocompromised host; CHD, congenital heart diseases; Chronic resp, other chronic respiratory conditions; yr, years. Analyses by χ^2^ test.

**Figure 2.**
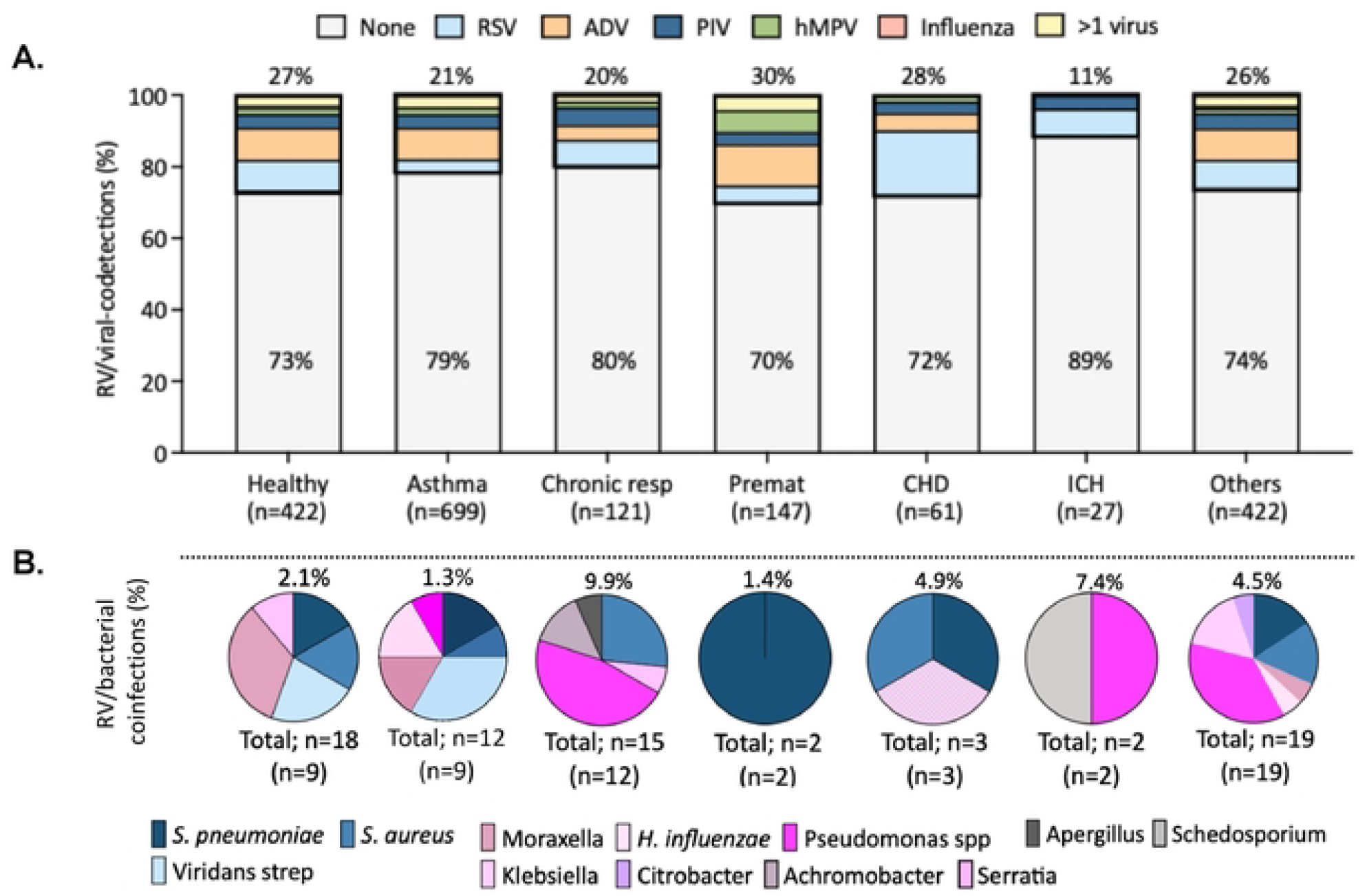
RV/viral co-detections and RV/bacterial coinfections in children and adolescents with RV infections. **A)** Bars represent the percentage of RV co-detection of each specific virus (RSV, ADV, PIV, hMPV, influenza) and those with co-detection of > 1 virus in the different groups. Percentage of RV-single infections are depicted inside the bars colored in grey, and percentage of RV/viral codetections are above each bar.

The prevalence of asthma doubled in children >1 year (45%-57%) *vs* infants (26%; p=0.007). Prematurity was more common in infants and children 1-5 years (10% and 7.6% respectively) than in older children (1.5%-2.5%; p=0.007). Only 0.2% of infants were ICH vs 4.6% of children 11 to <21 yr (p=0.007; **Table S2**). Upper respiratory infection (URI) was the most common diagnosis in ICH (55.6%) and healthy children (48.3%) while wheezing illnesses predominated in children with asthma (63.4%) and prematurity (49.7%). Pneumonia was the initial diagnosis in 42.2% of children with chronic respiratory conditions and in 18.5% of healthy children (p=0001). Fever, acute otitis media or sinusitis represented <4% of the admitting diagnosis.

In terms of laboratory data, 52.4% (995/1899) of children underwent blood work-up. Leukocytosis was more common in asthmatic children while leukopenia predominated in ICH. Abnormal radiologic findings were reported in 50% of ICH children *vs* 75%-85% of children with other comorbidities (p=0.017). The most common radiologic patterns were bronchial wall thickening and atelectasis in 56.0% to 67.2% of children (**Table S3)**.

### RV loads and Viral and bacterial co-infections

Semiquantitative RV loads (C_T_ values) were similar between healthy (25.6 C_T_ values) and children with comorbidities (25.05 C_T_ values), irrespective of the type of comorbidity (**Table S4**). A concomitant respiratory virus was identified in 26.5% (112/422) of healthy children and 19.8%-29.9% (347/1477) of those with comorbidities (**Fig 2A, Table S4**).

ADV and RSV followed by PIV were the most common co-detected viruses. RV/ADV were more common in asthmatic (8.9%) and premature children (11.6%; p=0.05), while RV/RSV co-detections were more frequent in CHD (18.0%), chronic respiratory conditions (7.4%) and children with other comorbidities (7.8%). In healthy children, rates of RV/ADV or RV/RSV co-detection were comparable and ∼9%. Co-detection of other respiratory viruses in ICH were rare and identified in only three children (RV/RSV, n=2; RV/PIV, n=1). More than one respiratory virus was identified in 2.8% (n=54) of children and the most frequent combination was RV/ADV/RSV in two-third of cases.

Bacterial co-infections were identified in 56 (2.9%) children based on lower respiratory/BAL cultures (n=48), CF (n=4) or blood cultures (n=4) and were more common in children with chronic respiratory conditions (9.9%) and ICH (7.4%); p=0.001; (**Table S4, Fig 2B)**. The most common bacteria were *S. pneumoniae, S. aureus and P. aeruginosa*.

**Figure 3. RV/viral co-detections and RV/bacterial coinfections in children and adolescents with RV infections. B)** Pie chart representing the proportion of different microorganisms isolated according to the different groups. The percentage of bacterial coinfections in each group is included above each pie chart, and the total number of bacteria and the number of patients (in parenthesis) is depicted underneath the pies. Pink and blue colors indicate gram-negative and gram-positive bacteria, respectively. Grey color indicates fungal pathogen. ADV, adenovirus; CHD, congenital heart diseases; hMPV, human Metapneumovirus; ICH, immunocompromised host; PIV, parainfluenza virus; Premat, premature; resp, respiratory; RSV, respiratory syncytial virus; RV, rhinovirus.

The proportion of gram-positive bacteria was greater in asthmatic children (58.3%) while gram-negative bacteria were mor commonly cultured in children with chronic respiratory conditions (71.4%). Differences did not reach statistical significance.

### Healthcare utilization in children with RV ARI

We assessed the use of medications and other indicators of healthcare utilization (**Table 2**). Systemic steroids were used more commonly in children with asthma, chronic respiratory conditions and prematurity (41.5-62.4%) than in healthy children (20.7%; *p*=0.0001). Bronchodilators including albuterol, ipratropium bromide or magnesium sulfate, were administered to ∼75% of children with asthma and other chronic respiratory diseases and in 14.8% of ICH, with no differences between groups (p>0.05). Antibiotics were administered to ∼40% of children, and their use was significantly higher in ICH (67%; *p*< 0.001).

**Table 2.** Healthcare utilization in children and adolescents with RV infection*.

|  | Previously healthy<br>(n=422) | Asthma/Atopy<br>(n=699) | Chronic respiratory conditions †<br>(n=121) | Prematurity<br>(n= 147) | CHD<br>(n=61) | ICH<br>(n=27) | Other §<br>(n=422) | p value |
| --- | --- | --- | --- | --- | --- | --- | --- | --- |
| **Medications** |  |  |  |  |  |  |  |  |
| **Systemic steroids, n (%)** | 87 (20.7) | 436 (62.4) | 58 (47.9) | 61 (41.5) | 21 (34.4) | 6 (22.2) | 90 (21.3) | **0.0001** |
| **Steroids duration (days)** | 2 [1-3] | 2 [1-3] | 3 [2-4] | 2 [1-3] | 3 [2-3] | 3.5 [2-8] | 2 [1-4] | **0.0019** |
| **Bronchodilators, n (%)** | 167 (39.6) | 535 (76.5) | 91 (75.2) | 94 (63.9) | 37 (60.7) | 4 (14.8) | 200 (47.4) | 0.65 |
| **Bronchodilator duration (days)** | 2 [1-4] | 1 [1-2] | 2 [1-4] | 2 [1-2] | 2 [1-7] | 2 [1-3] | 2 [1-4] | **0.0046** |
| **Antibiotics, n (%)** | 185 (43.8) | 246 (35.2) | 55 (45.5) | 63 (42.9) | 24 (39.3) | 18 (66.7) | 196 (46.4) | **0.0001** |
| **Duration (days)** | 5 [3-7] | 4 [2-7] | 5 [3-13] | 4 [2-7] | 8 [4-16] | 4.5 [3-11] | 5 [3-11] | 0.08 |
| **Hospital resources** |  |  |  |  |  |  |  |  |
| Hospital admission | 325 (77.0) | 614 (87.8) | 102 (84.3) | 132 (89.80) | 55 (90.2) | 22 (81.5) | 346 (81.9) | **0.0001** |
| Oxygen administration | 149 (35.3) | 456 (65.2) | 79 (65.3) | 84 (57.1) | 40 (65.6) | 5 (18.5) | 184 (43.6) | **0.0001** |
| PICU admission | 71 (16.8) | 300 (42.9) | 27 (22.3) | 49 (33.3) | 28 (45.9) | 3 (11.1) | 101 (23.9) | **0.0001** |
| Mechanical ventilation | 19 (4.5) | 37 (5.3) | 14 (11.6) | 16 (10.8) | 10 (16.4) | 1 (3.7) | 60 (14.2) | **0.0001** |
| Total hospital stay (days) | 1.75 [1.1-2.7] | 2.2 [1.4-3.6] | 3.0 [1.6-6.3] | 2.5 [1.4-4.3] | 3.2 [1.8-10.2] | 2.8 [1.1-5.2] | 2.5 [1.3-5.5] | **0.0001** |

\*Continuous variables are expressed as medians 25–75% interquartile ranges (IQR) and categorical data as frequency (%). Continuous variables were analyzed by Kruskal-Wallis test followed by Holms post-hoc test. Categorical data were analyzed by $\chi^2$ test. Numbers in bold indicate significant p values. CHD, congenital heart diseases; ICH, immunocompromised host; PICU, pediatric intensive care unit.
† Chronic respiratory conditions included: cystic fibrosis (n=18); chronic lung disease (CLD) (n=61); tracheoesophageal fistula, pulmonary hypertension, spontaneous pneumothorax, airway malacia, subglottic stenosis, cleft palate, and Pierre–Robin sequence. § Others included: genetic syndromes (n=132); gastrointestinal diseases (n=68); neurological disorders (n=36); hematologic disease (n=17) and varied including endocrine, osteoarticular or genitourinary underlying diseases (n=169).

Overall, 86% of children with comorbidities were hospitalized vs 77% of previously healthy (*p*=0.0001). The number of children who required supplemental oxygen was also higher in children with comorbidities (p<0.001). This difference was mainly driven by children with asthma (65.2%), CHD (65.6%) and other chronic respiratory diseases (65.3%) vs healthy (35%) or ICH (18.5%). PICU admission was higher in children with comorbidities (34%) vs healthy (17%) or ICH (11%); *p*=0.0001. This difference was also driven by children with asthma and CHD. Children with chronic respiratory diseases, CHD, prematurity or other underlying diseases required mechanical ventilation more commonly (11%-16%) than children with asthma, ICH or those previously healthy (<5%). Total length of stay was shorter for healthy children than those with comorbidities (1.7 vs 2.4 days; p=0.0001).

### Mortality and Predictor of Clinical Outcomes

Thirty children died, all but two had major comorbidities including asthma (n=7), ICH (n=4), CHD (n=2), chronic respiratory conditions (CLD; n=1), prematurity (n=1) and other comorbidities (genetic syndromes, n=11; neuromuscular diseases, n=1; central diabetes insipidus, n=1). Most of these children (63.3%; 19/30) had high RV loads (≤25 C_T_), intermediate (26-32 C_T_) in 23.3% (7/30), and low (>32 C_T_) in 4 (13.3%) children. A concomitant viral (n=6), bacterial (n=5) or fungal (n=1) co-infection was identified in 12 (40%) children. The per patient underlying condition, age, RV loads (ct values) and specific coinfection is included in **Table S5**.

Last, we performed multivariable analyses to define which comorbidity, adjusted by age, sex, RV loads and viral and bacterial coinfections, was associated with clinical outcomes defined as need for supplemental oxygen, PICU admission and longer length of stay, **Table 3**. Mortality was not included as an outcome due to the small sample size.

**Table 3.** Adjusted risk factors associated with RV disease severity.

|  | Supplemental O<sub>2</sub> |  | PICU |  | Length of stay |  |
| --- | --- | --- | --- | --- | --- | --- |
|  | OR (95% CI) | p-value | OR (95% CI) | p-value | OR (95% CI) | p-value |
| **Age** |  |  |  |  |  |  |
| Infants (<1yr) | **0.46 (0.31-0.68)** | **0.0001** | **0.43 (0.28-0.66)** | **0.0001** | **0.28 (0.18-0.44)** | **0.0001** |
| Toddlers (1-5yr) | 1.28 (0.85-1.93) | 0.233 | 1.23 (0.81-1.88) | 0.319 | **0.42 (0.27-0.65)** | **0.0001** |
| Children (>5-11yr) | 1.55 (0.98-2.47) | 0.060 | **1.66 (1.05-2.63)** | **0.029** | 0.65 (0.40-1.05) | 0.083 |
| **Sex (Female)** | 0.97 (0.79-1.18) | 0.775 | 0.99 (0.80-1.23) | 0.985 | 1.05 (0.86-1.28) | 0.626 |
| **Comorbidities** |  |  |  |  |  |  |
| Asthma/Atopy | **2.61 (2.00-3.41)** | **0.0001** | **2.82 (2.07-3.84)** | **0.0001** | **1.66 (1.26-2.19)** | **0.0001** |
| Chronic respiratory conditions † | **3.05 (1.96-4.75)** | **0.0001** | 0.97 (0.57-1.67) | 0.937 | **2.10 (1.34-3.28)** | **0.001** |
| Prematurity | **2.49 (1.68-3.71)** | **0.0001** | **2.52 (1.62-3.93)** | **0.0001** | **2.57 (1.72-3.86)** | **0.0001** |
| CHD | **3.00 (1.67-5.39)** | **0.0001** | **3.52 (1.95-6.35)** | **0.0001** | **2.62 (1.47-4.67)** | **0.001** |
| ICH | **0.22 (0.08-0.61)** | **0.004** | 0.31 (0.09-1.11) | 0.065 | 1.51 (0.64-3.53) | 0.341 |
| Others § | 1.24 (0.93-1.66) | 0.134 | 1.31 (0.92-1.86) | 0.133 | **1.96 (1.45-2.65)** | **0.0001** |
| **Viral co-detections** | 1.05 (0.84-1.33) | 0.760 | 1.16 (0.90-1.49) | 0.250 | 1.12 (0.89-1.42) | 0.309 |
| **Bacterial co-infections** | **2.06 (1.15-3.67)** | **0.001** | **5.43 (3.90-9.55)** | **0.0001** | **3.32 (1.77-6.22)** | **0.001** |
| **High RV loads (<25 Ct)** | 1.15 (0.94-1.41) | 0.162 | **1.40 (1.13-1.74)** | **0.002** | 1.12 (0.92-1.37) | 0.240 |

Odds ratios (ORs) and 95% confidence interval (CI). Numbers in bold indicate significant p values. For age, children >11 to 21 yrs was used as a reference. For sex, male was used as reference. For comorbidities, healthy was used as a reference. For viral co-detections and bacterial co-infections, none was used as a reference. For high viral loads, intermediate and low (>25-40 Ct) were used as a reference. O<sub>2</sub>, oxygen; PICU, pediatric intensive care unit. Length of stay was dichotomized by the median, and < 2.1 days used as a reference.
† Chronic respiratory conditions included: cystic fibrosis (n=18); chronic lung disease (CLD) (n=61); tracheoesophageal fistula, pulmonary hypertension, spontaneous pneumothorax, airway malacia, subglottic stenosis, cleft palate, and Pierre–Robin sequence. § Others included: genetic syndromes (n=132); gastrointestinal diseases (n=68); neurological disorders (n=36); hematologic disease (n=17) and other as endocrine, osteoarticular, and genitourinary underlying diseases (n=169).

Of all comorbidities, asthma, prematurity and CHD were consistently associated with increased odds of supplemental oxygen, PICU care and longer hospital stay. Chronic respiratory diseases were also independently associated with increase odds of oxygen supplementation and prolonged length of stay. Bacterial coinfections increased the risk of all three clinical outcomes, while older age and higher RV loads (<25 Ct values) were associated with increased odds of PICU care.

## DISCUSSION

In this study including ∼2,000 children and adolescents with RV ARI, we found that the majority had a chronic medical condition, and that the healthcare utilization, morbidity and even mortality was high and differed according to the type of comorbidity. Importantly, bacterial coinfections, older age and higher RV loads were also predictive of disease severity. To our knowledge this is the largest study conducted to date that analyzed the differential impact of symptomatic RV infections in children with existing comorbidities considering key factors that may have influenced clinical outcomes.

Studies have attributed lower rates of adverse outcomes among children with RV ARI likely due to the prolonged shedding of RV in the upper respiratory tract and the high rates of asymptomatic carriage ^4,5^. In our study, we found a high prevalence of underlying comorbidities in children with RV ARI, with asthma and prematurity being the most common, which has been previously described ^8,16-19^. We also found that the burden associated with RV ARI was high and that the need for hospitalization, supplemental oxygen, PICU admission or mechanical ventilation differed according to the type of underlying condition ^8,17,20-25^. While ICH had the lowest rate of healthcare utilization, asthma, prematurity and CHD were consistently and independently associated with worse disease severity ^10,26-28^. Differences between studies might be explained in part to the patients studied, as we only included children with symptomatic RV ARI ^16,29^. In addition, semiquantitative viral loads (C_T_ values) were available for all children, and median RV loads were overall intermediate-high suggesting the potential contribution of RV to the clinical phenotype. Older age was also associated with worse clinical outcomes, which contrasts with other respiratory viruses such as RSV, which is typically linked to increased disease severity in young children ^17,18,30^.

Studies have reported rates of viral codetection in children with RV ARI ranging from 15% to 50% depending on the study population and PCR platform used, a trend that has persisted post COVID-19 ^8,14,18,19,31,32^. We found that the rates of RV/viral co-detections were common and ranged from 20% to 30%. These proportions were consistent across the different types of underlying conditions except for ICH in whom RV/viral codetections were rare. As shown by others we also found that the most common co-detected viruses were ADV, RSV and PIV ^16,18,31^. Interestingly, the type of RV/viral codetection differed based on the underlying condition, with RV/ADV being more common in premature and asthmatic children, and RV/RSV in children with CHD and chronic respiratory diseases. Bacterial coinfections in children with RV ARI are not well described, likely due to the low prevalence of bacteremia in children with LRTI or that lower respiratory or pleural fluid samples are often unavailable, yet antibiotics are frequently used. Studies have reported antibiotic use in <30% of children with RV ARI, which is slightly lower than the 40% found in our study. This likely related to the higher prevalence of children with comorbidities that required hospitalization including ICH, that may have biassed clinical decision making ^25,33^. A case-control study of children with RV ARI found that bacterial coinfections were identified by sputum or throat cultures in ∼70% of inpatients vs 1% of outpatients ^19^. We identified bacterial coinfections in 2.9% of children, especially in ICH and those with chronic respiratory conditions. Our rates are similar to a study conducted in PICU children, that also defined bacterial coinfections according to lower respiratory tract or sterile site cultures ^32^. In our study, 1.5% of children died, and all but two had chronic medical conditions, with 40% having a viral or bacterial coinfection. Nevertheless, those and our study found that bacterial, but not viral, coinfections were consistently associated with worse clinical outcomes and warrant the inclusion of this parameter as possible contributor of RV disease severity in future studies.

This study has limitations. First, due to the retrospective study design data collection was limited to information recorded in the EHR. However, to address missing or incomplete data, we analyzed objective outcomes, and two independent investigators manually reviewed and curated the data to ensure accuracy. The patient population represented a convenience sample, which may be biassed to the more severe forms of the disease. The study however, included children evaluated in the outpatient setting, and although viral testing was performed at the discretion of the attending physician, it aligned with hospital policy during the study period. The viral panel did not include viruses like endemic coronaviruses or bocaviruses, which may have played a role on disease presentation and outcomes. We, however, we accounted for viral codetections as well as RV loads in multivariable analyses and did not find a significant effect of these factors on clinical outcomes. RV typing was not performed which limited our ability to assess the prevalence of RV-A, -B or -C infections in children with specific comorbidities and their contribution to clinical outcomes.

In summary, this study provides an in-depth characterization of the different comorbidities associated with severe RV infections in a large cohort of children and adolescents with RV ARI. It also highlights the important role that bacterial coinfections as well as age and RV loads have in RV-associated clinical outcomes. Defining the clinical phenotype of severe RV infections may help with resource allocation and patient triage, with the ultimate goal of improving clinical outcomes.

## Data Availability

All relevant data are within the manuscript and its Supporting Information files and summarized for publication purposes

## Notes

**Conflicts of Interest:** AM has received fees for participation in Advisory Boards from Janssen, Merck, Pfizer, Moderna, Enanta, AstraZeneca and Sanofi-Pasteur and grants from NIH, Merck and Janssen to institution. OR has received fees for participation in Advisory Boards from Merck, Pfizer, AstraZeneca and Sanofi-Pasteur and grants from NIH, Bill & Melinda Gates Foundation, Merck and Janssen to institution. ALL has received research grants from BioFire, Cepheid, Luminex, and Diasorin, and consulting fees from Medscape, Biorad and BioFire. The remaining authors have no conflicts of interest.

### Competing Interest Statement

I have read the journal's policy and the authors of this manuscript have the following competing interests: AM has received fees for participation in Advisory Boards from Janssen, Merck, Pfizer, Moderna, Enanta, AstraZeneca and Sanofi-Pasteur and grants from NIH, Merck and Janssen to institution. OR has received fees for participation in Advisory Boards from Merck, Pfizer, AstraZeneca and Sanofi-Pasteur and grants from NIH, Bill & Melinda Gates Foundation, Merck and Janssen to institution. ALL has received research grants from BioFire, Cepheid, Luminex, and Diasorin, and consulting fees from Medscape, Biorad and BioFire. The remaining authors have no conflicts of interest.

### Funding Statement

The author(s) received no specific funding for this work.

### Author Declarations

The study was approved by Nationwide Children’s Hospital (NCH) Institutional Review Board (IRB ID-STUDY00001015).

